# Assessment of Out of Pocket Expenditure and associated factors for availing COVID-19 vaccination by the beneficiaries in Bengaluru: South India

**DOI:** 10.1101/2022.01.29.22270032

**Authors:** Sunil Kumar Dodderi, H Lakshmi, J Srividya, S Manjula, R S Swathi

## Abstract

**Background:** Government of India has introduced COVID 19 vaccination in Jan 2021. There are no studies on out of pocket expenditure in COVID-19 vaccination in India, hence this study was undertaken to estimate the out of pocket expenditure for availing COVID 19 vaccine, to assess the factors associated with out of pocket expenditure for COVID vaccination and adverse events following immunisation.

**Methods:** This is a cross-sectional study conducted during Sep 2021-Dec 2021 of a medical college. A total of 438 study subjects above 18 years fulfilling inclusion and exclusion criteria were studied using probability proportional to population size. Data was collected using interview method by pre-tested semi structured proforma and analysed using descriptive & inferential statistics.

**Results:** The mean direct cost in Government vaccination centre was 3.24± 6.74 INR, indirect cost 809.10±1076.35 INR, total cost was 812.34 ±1079.49 INR.The mean direct cost in private vaccination centre was 1446.9±1845.65 INR, indirect cost 1140±1398 INR and total cost was 2586.90±2241.54 INR.

The mean total cost was OOPE for COVID 19 vaccination was 852.80 ±1128.512 INR, out of which direct cost was only 36.17(±359.20). The higher mean OOPE was found in loss of wages 670.02 INR. The factors associated with higher out of pocket expenditure was type of vaccine (P=0.031, OR=2.141, 95% CI=1.07-4.24) occupation of the study subject (P=0.000, OR=2.043, 95% CI= 1.37-3.03), reported stress following vaccination (P= 0.018, OR=1.72, 95%CI=1.098-2.703), adverse event within 48hrs (P=0.006, OR=2.125, 95% CI= 1.248-3.62), received any medication for adverse event (P=0.041, OR= 1.721, 95% CI= 1.022-2.84)

**Conclusion:** Majority of the study subjects utilized public facility. The higher mean out of pocket expenditure was for indirect cost loss of wages. This study shows that type of vaccine, occupation of the study subject and adverse event within 48 hrs, had 2 times higher out of pocket expenditure compared to other factors. Among the AEFI, fever was the most common, followed by pain at the injection site and myalgia.

## Introduction

Out of Pocket Expenditures (OOPE) are expenditures directly made by households at the point of receiving health care. This indicates the extent of financial protection available for households towards healthcare payments^1^ and is also described as health spending on hospitalization net of reimbursement ^2^.

In India, total health expenditure has been estimated to be 3.3% of gross domestic product and OOPE in health care services has been estimated to be 48.8% of the total health expenditure ^3^

The government’s efforts to improve public healthcare is conspicuous with out-of-pocket expenditure (OOPE) as a share of total health expenditure decreasing to 48.8% in 2017-18 from 64.2% in 2013-14^4^.

Around 90% of all households incurring impoverishing out-of-pocket health spending are already at or below the poverty line. Hence the need of an hour is to exempt the urban slum and poor people, from out-of-pocket health expenditure, with a better health financing policies.^5^

Out-of-pocket expenditure in health care includes traveling costs, loss of wages, cost of registration, consultation, hospital charges, and cost of medications, food and lodging of caregivers ^6^. Such out-of-pocket expenditures may lead to catastrophic health expenditures, where in the household has to borrow money or sell their property/assets or when they get contributions from friends/relatives to meet their health care expenses^7^

India began its largest COVID 19 vaccination drive on 16^th^ January 2021 to cover health care and frontline workers and scaled up to cover citizens above 45 years of age. The Government of India had selected the priority groups based on the potential availability of vaccines. Individuals with higher risk were given priority. From May 1^st^, 2021, all eligible citizens above the age of 18 years can get the COVID-19 vaccine.^8-10^

In India, Covishield (AstraZeneca’s vaccine manufactured by Serum Institute of India) in the month of January, Covaxin (manufactured by Bharat Biotech Limited) in the month of March, Sputnik -V in the month of April 2021, are the vaccines granted emergency use authorization by the Central Drugs Standard Control Organization^8-10^.

COVID 19 vaccination was provided both at the government and private vaccination centres^8^.

There is lack of data regarding OOPE in COVID 19 vaccination and it affects the economic stability of the household in India. When the expenditure on immunization affects the household, it affects the motivation of for vaccination and hence this research was undertaken and findings of first study on OOPE in COVID 19 vaccination in India will provide the hindsight.

## Objectives

1. To estimate the out of pocket expenditure for availing COVID-19 vaccination in adults above 18 years in Bengaluru, South India.
2. To assess the factors associated with out of pocket expenditure for availing COVID-19 vaccination in adults above 18 years in Bengaluru.
3. To describe the adverse events following immunization after availing COVID-19 vaccination in adults above 18 years in Bengaluru.

## Material and Methods

A cross sectional study was conducted in the urban field practice area, Department of Community Medicine of a medical college, Bengaluru from September 2021 – December 2021. Ethical Clearance was obtained from Institutional ethical committee, Akash Institute of Medical Sciences and Research Centre, Bengaluru, India. The study subjects included COVID -19 fully vaccinated individuals above 18 years of age. In September 2021, Karnataka has the coverage of fully COVID 19 vaccination of 26%, allowable error from prevalence is 20%, the sample size calculated was 273. Considering design effect of 1.5 and 10% additional size it was increased to 438. Respondents fully vaccinated for more than 15 days, who gave informed consent and resided for more than 6 months were enrolled.

The sampling method used is probability proportional to population size, population of urban poor locality was 7688, and number was 11. In each area, an approximate centre was marked, one of the roads was selected randomly by lottery method. After tossing coin one of the sides of road was chosen randomly. After walk through survey, using a currency note, a household was selected randomly. After interviewing a first household, the next household was selected by tossing the coin until the required sample is met.

The pre-tested tool consisted of both open and closed ended questions of the socio-demographic characteristics, type of vaccine, place of vaccination, side effects following vaccination and medications received. Expenditure incurred for COVID 19 vaccination was captured under two broad categories Direct cost (cost of vaccine, administration charges, cost of medicines, hospital charges) Indirect cost (transportation cost, expenses made for food, loss of wages of the individual and the accompanying person was obtained. A total of 438 study subjects above 18 years fulfilling inclusion and exclusion criteria were studied using probability proportional to population size.

The data collected was entered in MS excel and computed using SPSS 25. Descriptive statistics, inferential statistics logistic regression was used.

## Results

The mean age of the participants 42.37±14.41. The age range of the participants was 19 to 80. Table 1 describes the socio-demographic characteristics of the study participants.

**Table 1:** Socio-demographic profile of study participants.

| Age in years | Frequency(n=438) | Percentage (%) |
| --- | --- | --- |
| <20 | 17 | 3.9 |
| 21-30 | 99 | 22.6 |
| 31-40 | 94 | 21.5 |
| 41-50 | 101 | 23.1 |
| 51-60 | 80 | 18.3 |
| 61-70 | 40 | 9.1 |
| 71-80 | 7 | 1.6 |
| **Gender** |  |  |
| Male | 245 | 55.9 |
| Female | 193 | 44.1 |
| **Religion** |  |  |
| Hindu | 377 | 86.1 |
| Muslim | 61 | 13.9 |
| **Marital Status** |  |  |
| Married | 359 | 82.0 |
| Unmarried | 78 | 17.8 |
| Divorced | 1 | 0.2 |
| **Education Status of study subject** |  |  |
| Illiterate | 81 | 18.5 |
| Primary school | 35 | 8.0 |
| Middle school | 56 | 12.8 |
| High school | 121 | 27.6 |
| intermediate/diploma | 67 | 15.3 |
| graduate | 63 | 14.4 |
| professional/post graduate | 15 | 3.4 |
| **Occupation of study subject** |  |  |
| Unemployed | 163 | 37.2 |
| unskilled worker | 87 | 19.9 |
| Semi-skilled worker | 79 | 18.0 |
| Skilled | 37 | 8.4 |
| Clerical/shop Farmer | 37 | 8.4 |
| Semi-professional | 21 | 4.8 |
| Professional | 14 | 3.2 |
| **Education of Head of household** |  |  |
| Illiterate | 88 | 20.0 |
| Primary school | 52 | 11.9 |
| Middle school | 52 | 11.9 |
| High school | 138 | 31.5 |
| Intermediate/diploma | 60 | 13.7 |
| Graduate | 45 | 10.3 |
| Professional/post graduate | 3 | 0.7 |
| **Occupation of Head of household** |  |  |
| Unemployed | 7 | 1.6 |
| Unskilled worker | 42 | 9.6 |
| Semi-skilled worker | 112 | 25.6 |
| Skilled | 94 | 21.5 |
| Clerical/shop Farmer | 161 | 36.8 |
| Semi-professional | 12 | 2.7 |
| Professional | 10 | 2.3 |
| **Total income of Household** |  |  |
| <10000 | 80 | 18.3 |
| ≥10000 – 20,000 | 220 | 50.2 |
| ≥20000- 30,000 | 79 | 18.0 |
| ≥30000-40,000 | 30 | 6.8 |
| ≥40000-50,000 | 10 | 2.3 |
| 50,000> | 19 | 4.33 |
| **Co-morbidity** |  |  |
| Yes | 81 | 18.5 |
| No | 357 | 81.5 |

**Table 2:** Expenditure incurred for direct and indirect expenses for COVID 19 Vaccination.

| Direct cost (n=438) | Total Cost (INR) | Mean ± SD (INR) | Median (INR) | Range (INR) | 95% CI for Men | |
| --- | --- | --- | --- | --- | --- | --- |
|  |  |  |  |  | Lower Berth | Upper Berth |
| 1. Cost of vaccine | 13785 | 31.47(±352.35) | 0 | 0-5000 | 1.65 | 64.60 |
| 2. Medications | 2069 | 4.72(±41.86) | 0 | 0-500 | 0.79 | 8.65 |
| Total | 15854 | 36.17(±359.20) | 0 | 0-5000 | 2.42 | 69.96 |
| **Indirect cost** |  |  |  |  |  |  |
| 1.Travel expenses for the study participant and attendant | 64229 | 146.64(±262.04) | 100 | 0-2400 | 122.03 | 171.25 |
| 2. Loss of wages for the study participant and attendant | 293470 | 670.02(±1058.3) | 0 | 0-7500 | 570.63 | 769.41 |
| Total | 357699 | 816.66(±1083.81) | 400 | 0-7500 | 714.8 | 918.4 |
| **Grand Total** | **Rs. 373553** | 852.80±1128.512 | | | | |

\*Attendant loss of wages are taken for day of vaccination only (one day).

**Table 3:** Percentage distribution of the type, place, adverse event after receiving COVID-19 vaccination.

| Variable | N = 438 | Percentage (%) |
| --- | --- | --- |
| **Types of Vaccine** |  |  |
| Covishield | 398 | 90.9 |
| Covaxin | 40 | 9.1 |
| **Reported Stress following vaccination** |  |  |
| Yes | 335 | 76.5 |
| No | 103 | 23.5 |
| **AEFI within 48hrs** |  |  |
| Yes | 367 | 83.8 |
| No | 71 | 16.2 |
| **AEFI \*** |  |  |
| Fever | 265 | 60.50 |
| Myalgia | 161 | 36.8 |
| Pain at the injection site | 195 | 44.5 |
| Headache | 43 | 9.8 |
| Malaise | 13 | 3.0 |
| **Receive any medication for Adverse Events** |  |  |
| Yes | 367 | 85.6 |
| No | 71 | 14.4 |
| **Type of medicine** |  |  |
| Antipyretic | 325 | 74.2 |
| Analgesic | 32 | 7.3 |
| Antihistamine | 10 | 2.3 |
| None | 71 | 16.2 |

**Table 4:** Bivariate logistic regression of variables associated with OOPE on COVID 19 Vaccination.

| Variables in the Equation |  |  |  |  |  |  |  |
| --- | --- | --- | --- | --- | --- | --- | --- |
|  | B | S.E. | Wald | df | Sig. | Exp(B) | 95% of CI<br>LB-UB |

|  |  | Co-<br>efficient<br>constant |  |  |  |  |  |  |
| --- | --- | --- | --- | --- | --- | --- | --- | --- |
| Step 1<sup>a</sup> | Age | -.006 | .007 | .832 | 1 | .362 | .994 | .981 -1.07 |
|  | Constant | .221 | .298 | .550 | 1 | .458 | 1.247 |  |
| Step 1<sup>a</sup> | Gender (1) | .437 | .194 | 5.085 | 1 | .024 | 1.548 | 0.442 -0.944 |
|  | Constant | -.282 | .145 | 3.752 | 1 | .053 | .755 |  |
| Step 1<sup>a</sup> | Comorbidity | .229 | .247 | .855 | 1 | .355 | 1.257 | 0.774 -2.401 |
|  | Constant | -.452 | .460 | .967 | 1 | .325 | .636 |  |
| Step 1<sup>a</sup> | Type of vaccine | .761 | .352 | 4.670 | 1 | **.031** | **2.141** | **1.073 -4.24** |
|  | Constant | -1.492 | .683 | 4.778 | 1 | .029 | .225 |  |
| Step 1<sup>a</sup> | Place of vaccination | 1.452 | .796 | 3.322 | 1 | .068 | 4.271 | 0.892 -20.34 |
|  | Constant | -1.517 | .814 | 3.475 | 1 | .062 | .219 |  |
| Step 1<sup>a</sup> | Education | .290 | .248 | 1.368 | 1 | .242 | 1.337 | 0.94 -2.65 |
|  | Constant | -.563 | .461 | 1.494 | 1 | .222 | .569 |  |
| Step 1<sup>a</sup> | Occupation | .714 | .202 | 12.521 | 1 | **.000** | **2.043** | **1.37-3.035** |
|  | Constant | -1.202 | .345 | 12.164 | 1 | .000 | .300 |  |
| Step 1<sup>a</sup> | Education of household | -.074 | .240 | .096 | 1 | .756 | .928 | 0.581 -1.45 |
|  | Constant | .097 | .442 | .048 | 1 | .826 | 1.102 |  |
| Step 1<sup>a</sup> | Occupation of head of household | 21.198 | 15191.270 | .000 | 1 | .999 | 1607969059.351 | 0.000 |
|  | Constant | -42.401 | 30382.541 | .000 | 1 | .999 | .000 |  |
| Step 1<sup>a</sup> | Duration | .009 | .007 | 1.618 | 1 | .203 | 1.009 | .995 -1.02 |
|  | Constant | -.163 | .137 | 1.399 | 1 | .237 | .850 |  |
| Step 1<sup>a</sup> | Reported stress following vaccination | .544 | .230 | 5.598 | 1 | **.018** | **1.723** | **1.098-2.703** |
|  | Constant | -.998 | .419 | 5.678 | 1 | .017 | .369 |  |
| Step 1<sup>a</sup> | Adverse event within 48 hr of vaccination | .754 | .272 | 7.695 | 1 | **.006** | **2.125** | **1.248-3.62** |
|  | Constant | -1.426 | .513 | 7.741 | 1 | .005 | .240 |  |
| Step 1<sup>a</sup> | Received any medication for side effect | .543 | .266 | 4.171 | 1 | **.041** | **1.721** | **1.022 -2.84** |
|  | Constant | -1.031 | .500 | 4.255 | 1 | .039 | .357 |  |
| a. Variable(s) entered on step 1: Gender, Comorbidity, Type of vaccine, Place of Vaccine, Education, Occupation, Education of household, Occupation of Household, Duration, Side effect, reported stress following vaccination, Received and medication for side effect |  |  |  |  |  |  |  |  |

**Table 5:** Predicted mean expenditure (in rupees) by socio-demographic factors and place of vaccination, type of vaccination (Total expenditure in rs)

| Parameter Estimates |  |  |  |  |  |  |  |
| --- | --- | --- | --- | --- | --- | --- | --- |
| VAR00009<sup>a</sup> | B | Std. Error | Wald | df | Sig. | Exp(B) | 95% Confidence Interval for Exp(B) |

|  |  |  |  |  |  |  |  | Lower Bound | Upper Bound |
| --- | --- | --- | --- | --- | --- | --- | --- | --- | --- |
| 1.00 | Intercept | -4.374 | 1.788 | 5.986 | 1 | .014 |  |  |  |
|  | Age | -.022 | .011 | 4.034 | 1 | .045 | .979 | .958 | .999 |
|  | Gender | -.516 | .230 | 5.012 | 1 | .025 | .597 | .380 | .938 |
|  | Education | -.313 | .096 | 10.530 | 1 | .001 | .731 | .606 | .884 |
|  | Occupation | .096 | .072 | 1.743 | 1 | .187 | 1.100 | .955 | 1.268 |
|  | Education of head of household | -.027 | .073 | .132 | 1 | .716 | .974 | .844 | 1.124 |
|  | Total income of household | .000 | .000 | .008 | 1 | .931 | 1.000 | 1.000 | 1.000 |
|  | Comorbidity | .045 | .291 | .024 | 1 | .877 | 1.046 | .591 | 1.850 |
|  | Duration | .009 | .008 | 1.236 | 1 | .266 | 1.009 | .993 | 1.024 |
|  | Type of vaccine | 1.037 | .379 | 7.481 | 1 | **.006** | **2.820** | **1.342** | **5.928** |
|  | Place of vaccination | 1.643 | .876 | 3.520 | 1 | .061 | 5.172 | .929 | 28.790 |
|  | Reported stress following vaccination | .183 | .363 | .255 | 1 | .614 | 1.201 | .590 | 2.446 |
|  | AEFI within 48hr of vaccination | .789 | .514 | 2.360 | 1 | .125 | 2.202 | .804 | 6.028 |
|  | Received any medication for AEFI | -.200 | .419 | .228 | 1 | .633 | .819 | .361 | 1.860 |
| a. The reference category is: .00. |  |  |  |  |  |  |  |  |  |

Majority of the study participants had hypertension 52 (64.2%), followed by diabetes mellitus 35 (43.2%) and thyroid disorders 7 (8.6%).

Majority of study participants received the vaccination at the 1^st^ visit at the vaccination centre 426(97.3%), 2nd time 11 (2.5%), 3^rd^ time 1 (0.2%). Participants were accompanied by a family member 281(64.1%), and used personal mode of transport 394 (90.0%). Mean duration of time to the health facility 14.74±14.20, Range (90-0)

**Graph 1:**
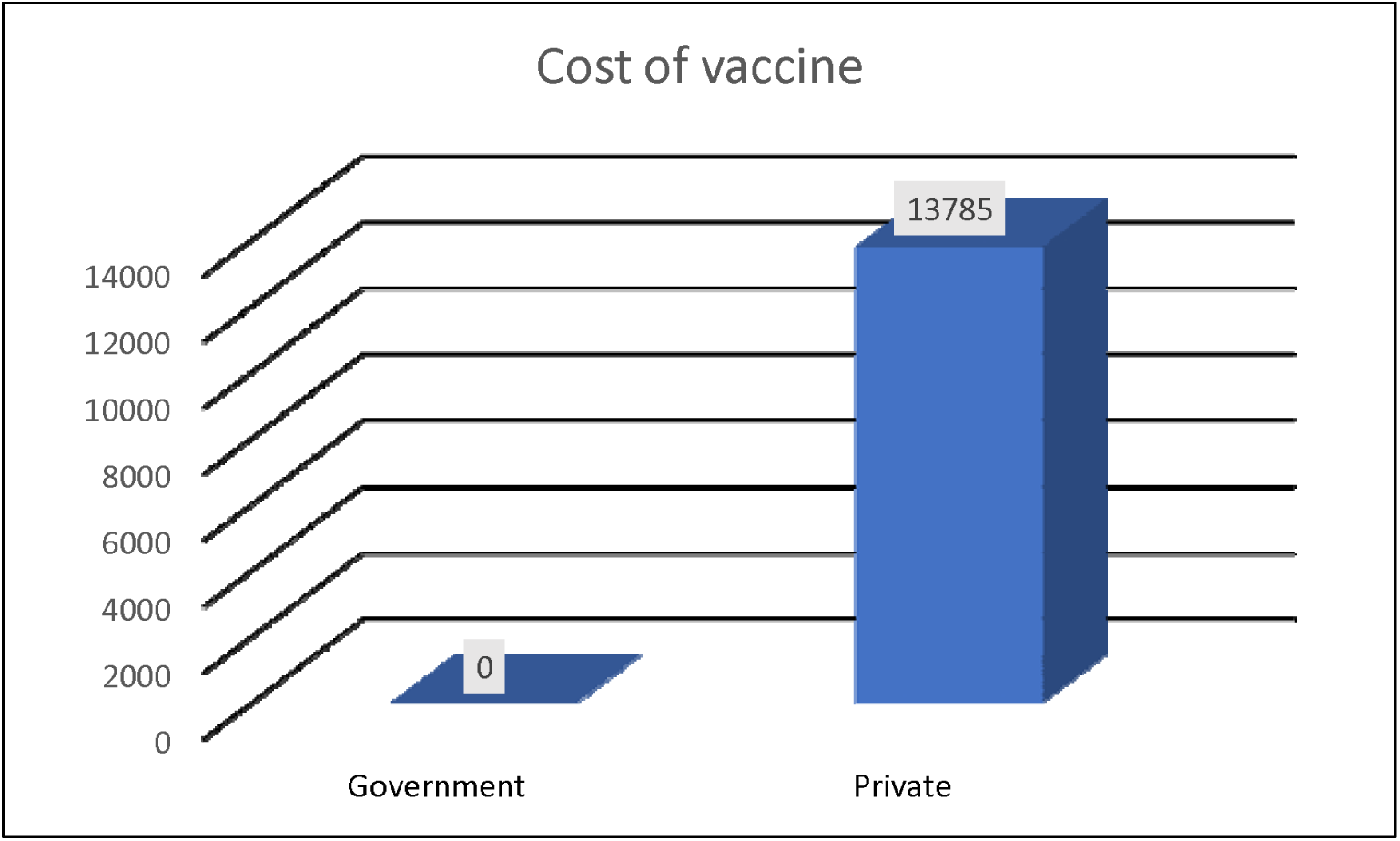
Distribution of study participant for cost of vaccine in Government and Private Vaccination Centres.

The cost of vaccine included the hospital charges in government facility it was 0 INR. The average cost of vaccine in private hospital was 13785 INR.

**Graph 2:**
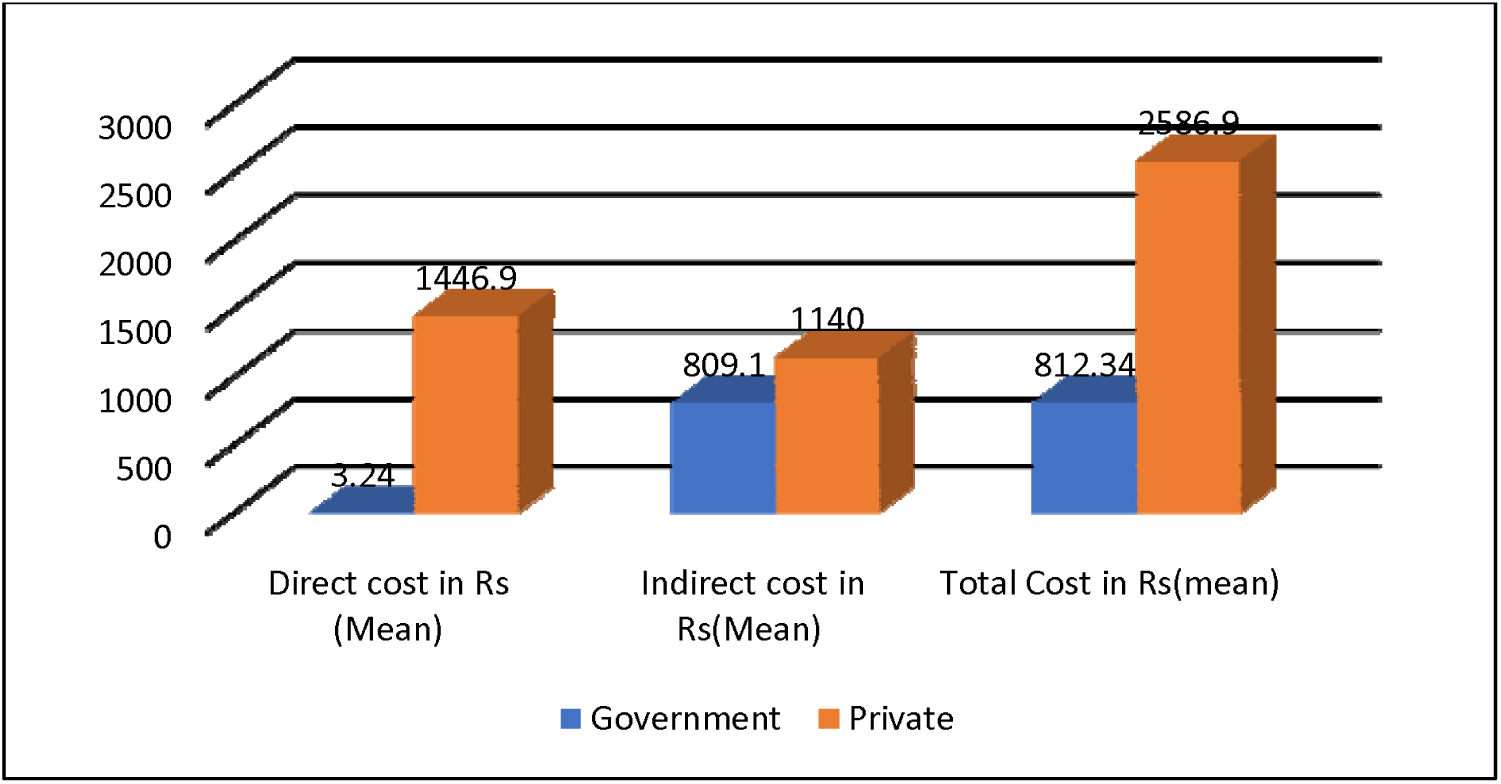
Distribution of study participants according to cost incurred in Government and Private Vaccination Centers.

The mean direct cost in Government vaccination centre is 3.24± 6.74 INR, indirect cost 809.10±1076.35 INR, total cost 812.34 ±1079.49 INR, Median 400, Range 0-1200 INR, 95%

CI 128.39-801.60.

The mean direct cost in private vaccination centre is 1446.9±1845.65 INR, indirect cost 1140±1398 INR and total cost 2586.90±2241.54 INR, median 1970, range 0-5800, 95% CI 983.39-4190.40.

To assess the factors associated with out of pocket expenditure in study participants, age, gender, co-morbidity, type of vaccine, place of vaccine, education of the study participant, occupation of the study participant, education of head of household, occupation of head of household, time duration to vaccination centre, reported stress following vaccination, adverse event within 48hrs, received any medication for adverse event were included in binary logistic regression model. The method used was the forward regression model.

In this study it was intended to assess the factors associated with higher out of pocket expenditure as an outcome variable. Therefore, higher out of pocket expenditure total cost ≥ 500 is coded as 1, < 500 as coded as 0 as a dependant variable. The other factors listed earlier which was basically categorical was coded 0 and 1.

The factors gender, type of vaccine,reported stress following vaccination, received any medication for adverse event showed statistically significant results P<0.05. The factors occupation of study subject, adverse event within 48 hrs showed highly significant results P<0.01.

However, the factors associated with higher out of pocket expenditure was type of vaccine (P=0.031, OR=2.141, 95% CI=1.07-4.24) occupation of the study subject (P=0.000, OR=2.043, 95% CI= 1.37-3.03), reported stress following vaccination (P= 0.018, OR=1.72, 95%CI=1.098-2.703), adverse event within 48hrs (P=0.006, OR=2.125, 95% CI= 1.248-3.62), received any medication for adverse event (P=0.041, OR= 1.721, 95% CI= 1.022-2.84). This study shows this type of vaccine, occupation of the study subject and adverse event within 48 hrs, had 2 times higher out of pocket expenditure with the Odds Ratio of 2.14, 2.04 and 2.12 respectively.

The factors which were included in the bivariate logistic regression was included in the multivariate logistic regression and the factor type of vaccine P value 0.006, was highly significant, with Odds Ratio =2.820, 95% CI=1.342-5.928 and was associated with higher out of pocket expenditure and rest of the factors were not significant statistically.

## Discussion

The COVID⍰19 pandemic caused nearly unprecedented harm to many nations’ lives, health, and economies. Since Vaccination is an effective measure to prevent against infectious diseases COVID⍰19 vaccination prevent the severity of the illness and have played a vital role in preventing the mortality.

Out-of-pocket expenditure is a major problem. The National Health Policy 2017^12^ estimated that 7% of the Indian population is pushed into poverty each year because they are not able to afford the OOP costs.

In the current study majority of the study subjects utilized the public facility 97.7%, for COVID 19 vaccination, may be explained due to the study place which was in urban poor locality.

Under the National COVID Vaccination Programme, the Government Vaccination Centres provided free of cost to all the eligible beneficiaries. In the Private Vaccination Centres cost of vaccines was on higher side, so the Centre capped the maximum price, for Covishield 780 INR, Covaxin 1410 INR, Sputnik V 1145 INR in June 2021^13^.

In the current study cost of vaccine in government facility was 0 INR which implies successful implementation of COVID 19 vaccination drive in India. In order to pace the vaccination programme and individuals who can afford the government had made availability of vaccine in the private hospital, where average cost of vaccine in private was 1378.5 INR per person which was on a higher side.

According to this study higher mean out of pocket expenditure was incurred for indirect cost i.e., loss of wages 670.02 INR. This study is in contrast to observation study conducted on Yellow fever immunization^14^ and Routine Immunization^15^ where travel expenses cost was higher.

Study found out the higher mean out of pocket expenditure was for indirect cost loss of wages followed by travel expenses, this has to be lowered as this may act as reasons for vaccine hesitancy. The factors associated with higher OOPE was 2 times in type of vaccine, occupation of the study subject and adverse event within 48hrs.

To the best of our knowledge this is the only study calculated on out of pocket expenditure on COVID 19 vaccination in adults. Limitation of the study could be the recall bias of the study participants.

With less than a third of health expenditure covered by the government, the major burden of India’s health costs falls on individual households, per the <u>National Health Account Estimates</u>. Household out-of-pocket expenditure on health made up for 58.7% of the total health expenditure, <u>One in five</u> Indians fall below the poverty line and more are pushed into poverty every year due to health expenses, according to World Bank data.

The central government’s expanded Covid-19 vaccination procurement policy also allows private hospitals to purchase doses directly from manufacturers, and to set prices for consumers. Currently, both SII and Bharat Biotech are providing doses to private hospitals at higher rates than to state governments. SII set the rate of Covishield for private hospitals at Rs 600 per dose and Bharat Biotech set the rate at Rs 1,200. Given this policy, manufacturers have a financial incentive to divert more stocks towards private hospitals. The entire course of two doses for people getting vaccinated in private facilities is a minimum Rs 1,200-Rs 2,400, just for the doses. Private hospitals also add <u>charges</u> for administering the vaccines.

These rates are unaffordable for a large section of the Indian population. The monthly per capita income of an average four-member household was Rs 4,979 in October 2020, seven months after the Covid-19 nationwide lockdown was imposed – a 17% decrease from the level of Rs 5,989 in January 2020, per Azim Premji University’s <u>State of Working Report India 2021</u>, published this month. The number of individuals who earn less than the national minimum wage threshold (<u>Rs 375 per day</u>) increased by 23 crore during the pandemic, said the report.

Our study revealed the most common adverse event was fever in 60.50%, myalgia in 36.8%, pain at the injection site 44.5%, headache in 9.8%and malaise in 3.0 %. which was in contrast Study conducted in Bangladesh by Parvej^16^ et al showed fever (36.05%), muscle pain (31.69%), pain in the injection site (30.67%), and headache (23.40%) can be due to different geographical location. Around 85.6 % had received medication for the adverse events, which was in contrast to study conducted by Parvej et al^16^ where only 37.65 had received medication. In the current study the most common medication received is antipyretic 74.2%, which is similar Jain^17^ et al. Majority of the study subjects had hypertension 64.2%, where has study conducted on health workers^17^ only 4.5% had hypertension.

## Conclusion and Recommendations

Majority of the study subjects utilized public facility. The higher mean out of pocket expenditure was for indirect cost loss of wages followed by travel expenses. However the factors associated with higher out of pocket expenditure was type of vaccine, occupation of the study subject, reported stress following vaccination, adverse event within 48hrs, received any medication for adverse event.

The out-of-pocket expenditure is one of the hidden expenditures which is not included in the vaccination cost during immunization. To conclude the COVID -19 vaccination has proved to have the out-of-pocket expenditure of considerable amount, which is in the form of area of residence, traveling cost, traveling distance, and indirectly loss of wages due to time spent in traveling, waiting, and process of vaccination. Through this study, we could conclude that it could be one of the reasons for lower vaccination coverage, and vaccine hesitancy can be associated with this extra expenditure. The health providers are given allowances for vaccination services but still, policymakers should take ‘out-of-pocket expenditure’ of the families of vaccinee into consideration because it may lead to catastrophic health expenses shattering the economy of the recipient family. Our government has already started health insurance benefits for a large segment of the population, and out-of-pocket expenditure due to immunization should be included under that initiative. The traveling cost can be reduced by providing quality services in the periphery.

In a country like India where social security schemes are not universal, out of pocket expenditure for vaccination may bring financial stress to the family and maybe an important reason to delay or deny vaccination. Social and health insurance policies should cover immunization as it is an important preventive strategy to combat vaccine-preventable communicable diseases.

Reducing out of pocket expenditure can involve program and policy changes that make vaccinations more affordable. Changes could include paying for vaccination administration, providing new or expanded insurance coverage (copayments / co-insurance).

COVID 19 vaccines are safe and the adverse events are mild. All the adults have to be immunized to fight against the COVID-19 deadly disease.

## Data Availability

All data produced in the present study are available upon reasonable request to the authors

## Acknowledgement

Authors would like to thank Postgraduate Dr.Lavanya.K, MBBS interns Ambika, Anjaly K Prakash, Anusha G V, Ashwini A, Bhoomika Choudary, Bhomika M O and Medical social workers Akshay Kumar, Shivaprasad and Durgesh for the support rendered for the smooth conduction of study. Authors are grateful for the study participants.

## Ethical approval

The study was approved by the Institutional Ethical Committee, Akash Institute of Medical Sciences and Research Centre, Bengaluru, India.

## Conflict of interest

None declared

## Funding

No funding sources.

## Notes

### Competing Interest Statement

The authors have declared no competing interest.

### Author Declarations

Institutional ethical committee , Akash Institute of Medical Sciences and Research Centre, Bengaluru, India.

